# Analysis of the Interventions Adopted Due to the COVID-19 on Ari Morbility for Colombia

**DOI:** 10.1101/2020.09.12.20193334

**Authors:** Á. Quijano-Angarita, O. Espinosa, M. Mercado, D. Walteros, D. Malo

**Affiliations:** Instituto de Evaluación Tecnológica en Salud (IETS), Bogotá, Colombia; Demographic and Epidemiological Observatory of the Andean Area (ODEAN), Universidad Nacional de Colombia, Bogotá, Colombia; Research group on Economic Models and Quantitative Methods, Universidad Nacional de Colombia, Bogotá, Colombia; Instituto Nacional de Salud – INS. Bogotá, Colombia

## Abstract

Acute Respiratory Infections are among the leading causes of death globally, particularly in developing countries, and are highly correlated with the quality of health and surveillance systems and effective early interventions in high-risk age groups. According to the World Health Organization, about four million people die each year from mostly preventable respiratory tract infections, making it a public health concern. The official declaration of a pandemic in March 2020 due to the Sars-CoV-2 virus coincided with the influenza season in Colombia and with environmental alerts about low air quality that increase its incidence. The objective of this document is the application of a flexible model for the identification of the pattern and monitoring of ARI morbility for Colombia by age group that shows atypical patterns in the reported series for 5 departments and that coincide with the decisions implemented to contain the COVID-19.

## Introduction

Acute Respiratory Infection (ARI), defined as a set of infections caused by viral and bacterial microorganisms to the respiratory system of high dissemination, presents the highest morbidity in the world and is among the leading causes of medical care and death in children under 5 years in Colombia, with an accentuated pattern in certain seasons, in the form of epidemic outbreaks, which vary according to the climatic and epidemiological characteristics of the regions. Cases of acute respiratory infection include clinical diagnoses that involve everything from common colds to more serious cases that require, as in the case of pneumonia, specialized hospital care that includes managing patients in hospitalization or intensive care areas because they are a threat to life.

According to the WHO (1), lower respiratory tract infections cause more than four million deaths per year at high rates in low- and middle-income countries and in children under 5 years of age. In fact, in 2015 for the 0-5 age group, respiratory infections caused 920.136 deaths, representing 15% of deaths among all causes worldwide. In Colombia, according to data calculated from vital statistics report (Table 1 from National Statistics Department of Colombia - DANE data), in 2017 and 2018 deaths from diseases in the respiratory system in children under five years of age represented 8.3% and 8.5% respectively of the total number of all causes of death in this age group, where between 5.6% and 5.8% of deaths were caused by influenza. As the cases due to ARI represent a high risk of death and, in addition, a high demand for medical assistance, they represent an event of interest in public health also because attendance at health services leads to treatment failure and high risk of death.

**Table 1.** Deaths caused by respiratory diseases and percentage of total causes of death in children under-five, Colombia 2017–2018.

| CAUSES OF DEATHS | 2017 |  | 2018 |  |
| --- | --- | --- | --- | --- |
|  | Deaths | total %<br>causes | Deaths | total %<br>causes |
| Pneumonia | 500 | 5.84% | 508 | 5.7% |
| Lung diseases due to external agents | 12 | 0.14% | 9 | 0.10% |
| Chronic diseases of the lower respiratory tract | 12 | 0.14% | 21 | 0.23% |
| Other diseases of the respiratory system | 184 | 2.15% | 221 | 2.47% |
| Other causes of death | 7840 | 91.71% | 8174 | 91.5% |

**Source:** own calculations based on vital statistics (DANE).

The WHO declaration of a global pandemic (March 11, 2020) due to the novel coronavirus COVID-19 and the appearance of the first case imported into Colombia (March 6) from Italy of this easily transmitted infection due to the Sars-CoV-2 virus, coincides with the beginning of the influenza season in the country, the air pollution alerts in the main cities of the country such as Bogotá and Medellín and the humanitarian crisis due to the massive migration of Venezuelan citizens that, is expected to have an impact on the number of cases reported of Acute Respiratory Infection, and therefore, that there is concern about the hospital capacity and the pressure this could cause in the Colombian health system (leading to a collapse in the hospital network causing an increase in deaths from preventable causes). On the contrary, that because of the epidemic and the measures taken the peaks of IRA morbility fall and capacity can be released.

The lockdown decreed by the Colombian government on March 24, and extended until April 27 throughout the country, was defined after the measure to close schools and universities and cancel massive events (March 11 and 12, 2020) and was intended to reduce the speed of contagion from Sars-CoV-2 and avoid a possible collapse of health systems that in the Latin American region are quite fragile (2). Therefore, the assessment and monitoring of health events, especially those related to respiratory diseases, are important, especially for different age groups and for outpatient and serious cases involving hospitalization.

Therefore, this paper first presents an analysis of the number of cases adjusted by the population and a Bayesian model of binomial negative response, to determine the departmental pattern of ARI morbility in outpatient (including medical emergencies) and inpatient (including medical intensive care) by age groups (0-4, 5-19, 20-59, 60+ years) to enable the evaluation of interventions implemented at the national level to contain COVID-19 and decision-making in prevention and control.

## Materials and methods

### Data

For the analysis, we used the database of Sivigila (Sistema Nacional de Vigilancia en Salud Pública) of event 995 - Morbility due to ARI of the Instituto Nacional de Salud – INS (National Institute of Health), which contained information on counts of Acute Respiratory Infection outpatient and emergency, hospitalization and critical care by departments and large cities and for the 52 epidemiological weeks of the year, in addition to information on other causes of disease outpatient, emergency and hospitalization (including critical care). The database was processed from the first week of 2017 to week 13 of the year 2020 for risk calculation at the departmental level and for Bayesian model (time interval with the best quality records). In addition, the estimated and projected population presented by the National Administrative Department of Statistics (DANE) of Colombia was obtained to calculate the population at risk by using an exponential interpolation per weeks, using official population data between 2018 and 2021. The data was processed by department, merging large cities data with the department to which the belong to produce an aggregate analysis.

### Method

First, for an exploratory analysis the risk is calculated at a departmental level per week, defined as,

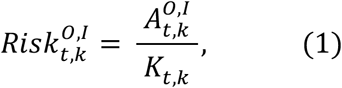

where risk (1) is expressed as the quotient between the new ARI cases (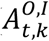) per week t and department k observed for outpatients (O) and inpatients (I) and the population at risk (*K_t,k_*), which was obtained at a weekly level by using exponential interpolation defined as: *K_o_* exp(*rt*) where r is the weekly growth rate, and *K_o_* is the initial population.

Second, since we have cases of new reported cases of ARI morbility, we use a negative binomial model as simple as possible that considers the variability and dependence between epidemic weeks and one that allows for dealing with the apparent over-dispersion present in the counts of some departments of Colombia due to the quality of the reported data. Let *y_i_* ∼*Negbin*(*p_i_, r*) be the number of cases of ARI morbility by weekly (from week 1 of 2017 to week 13 of 2020), age group and for inpatients and outpatients, with *i* = 1,…, 169, we use the parameterization presented by Ntzoufras3 whose density function is defined as,

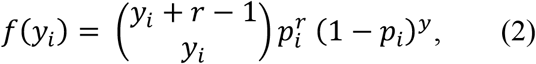

where 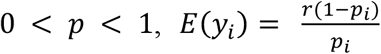 and 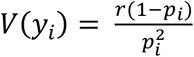. Then, we model the interest variable *y_i_* from a Bayesian model defining parameters from (2) as follows,

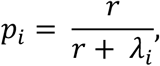

with,

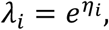

and the predictor lineal *η_i_*, considering year and week variables as covariates,

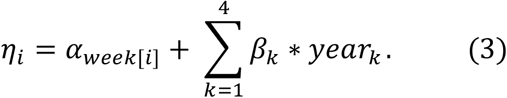

In (3) we considered an intercept that varies for each epidemiological week (1-52) and considers the dependence between them and a linear combination of dummy variables for each year (2017-2020) with,

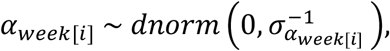

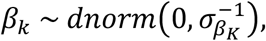

and non-informative priors for the standard deviations such that,

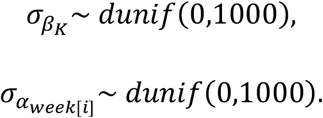

The model was fitted using JAGS, simulating 3 chains of 30,000 draws each with a burning period of 15,000. To evaluate convergence, the Gelman-Rubin convergence diagnostics is followed (4) and is considered a threshold of 1.1 as suggested by Gelman and Rubin (5).

The estimation accuracy is measured by the symmetric Mean Absolute Percentage Error (sMAPE), which has the advantage of avoiding the asymmetry of MAPE and is less sensitive to outliers and satisfy 0 < sMAPE < 1 (by dividing by 2 the original sMAPE which considers an arithmetic mean in the denominator). Thus, the sMAPE used is defined as follows^1^,

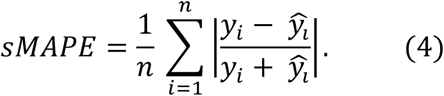

### Results and discussion

Figures 1-5 show the calculated risk for the departments of Bogotá, Antioquia, Boyacá, Huila and Valle del Cauca with atypical behavior in the last weeks of monitoring^2^. From a descriptive point of view, the series show a notable change in cases of Acute Respiratory Infection - ARI since week 10, when the first case was reported. The trend observed is an exaggerated increase between week 10 and 11 of outpatient cases in the 20-59 age groups for the 5 departments, cases that decrease by week 11 when it is decided to close schools, universities and ban massive events in the country. A striking behavior in the series is that in the department of Valle del Cauca for the 5 to 19 year age group where the number of cases of patients in hospitalization increased with respect to previous years. Between week 11 and 13, when the lockdown is decreed and children over 70 are prohibited from commute, the series falls below the level of previous years, an effect that is perceived in all departments. By week 13 in the department of Huila, there is an increase in the number of cases of hospitalization in the 5-19 age group, and their evolution must be monitored.

**Figure 1.**
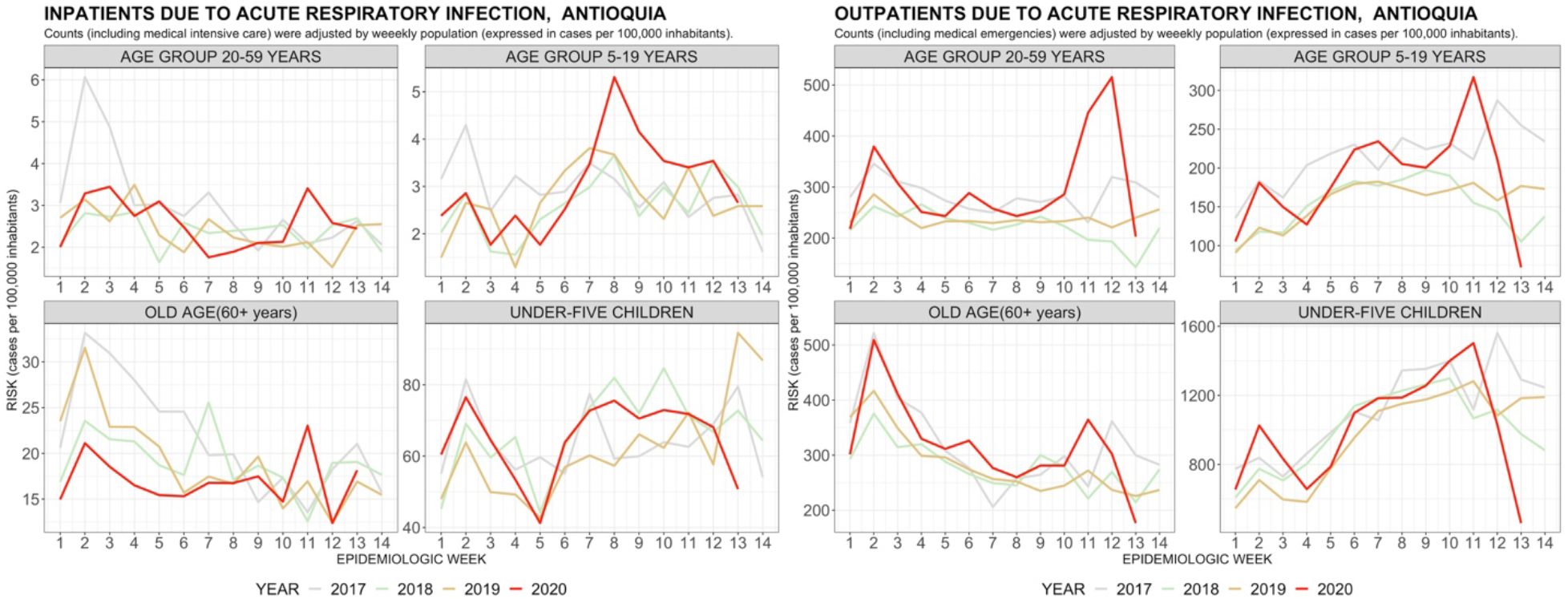
Direct estimation of risk due to Acute Respiratory Infection – ARI, Antioquia.

**Source:** own calculations.

**Figure 2.**
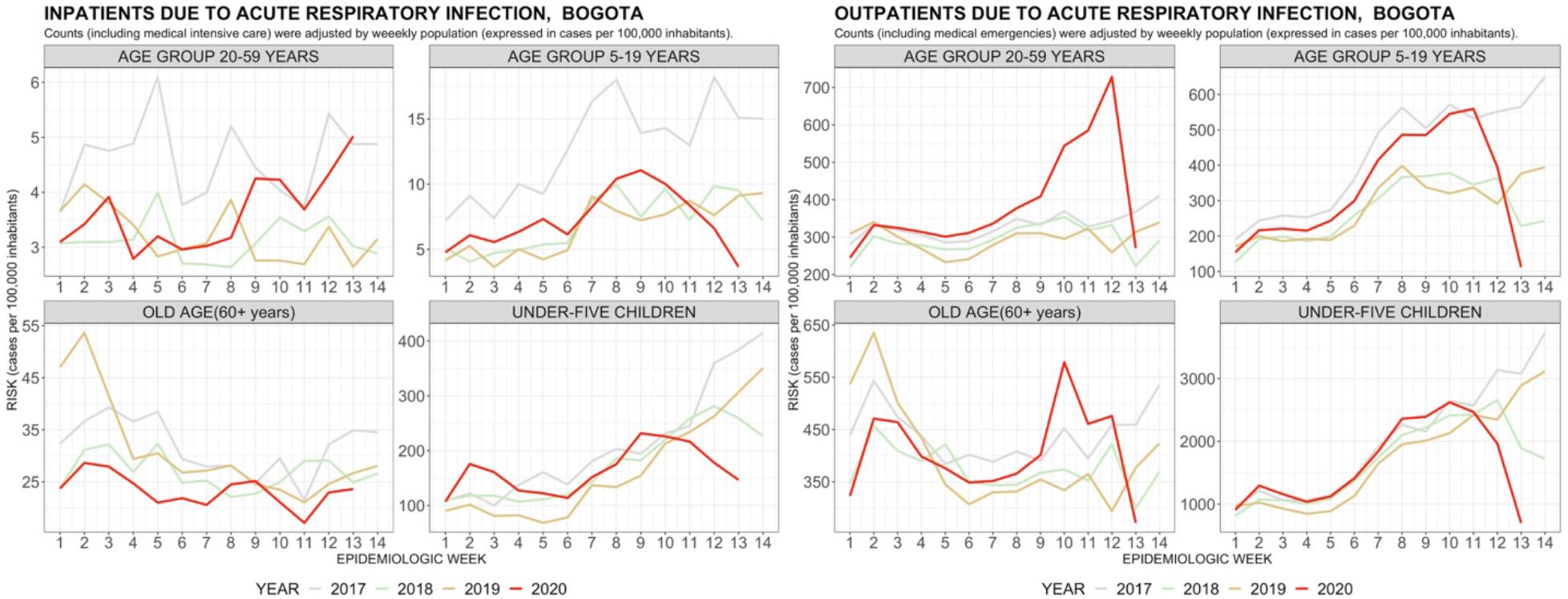
Direct estimation of risk due to Acute Respiratory Infection – ARI, Bogotá.

**Source:** own calculations.

**Figure 3.**
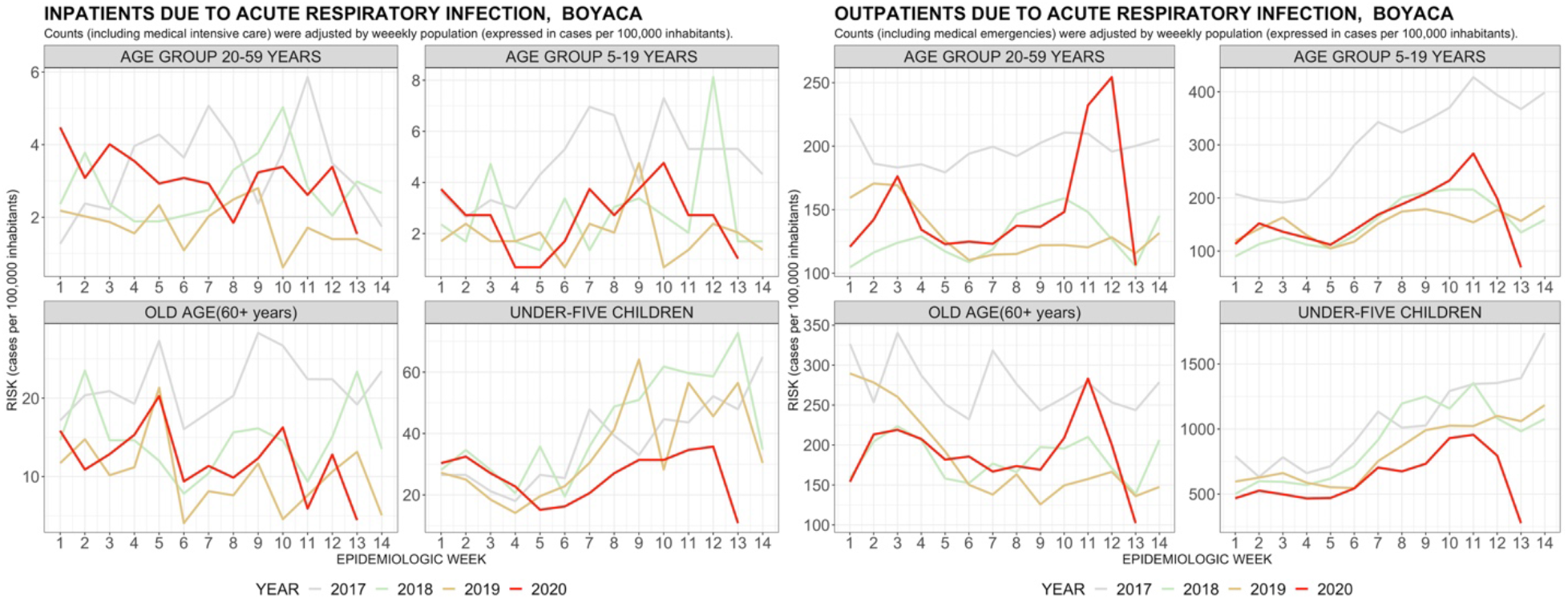
Direct estimation of risk due to Acute Respiratory Infection – ARI, Boyacá.

**Source:** own calculations.

**Figure 4.**
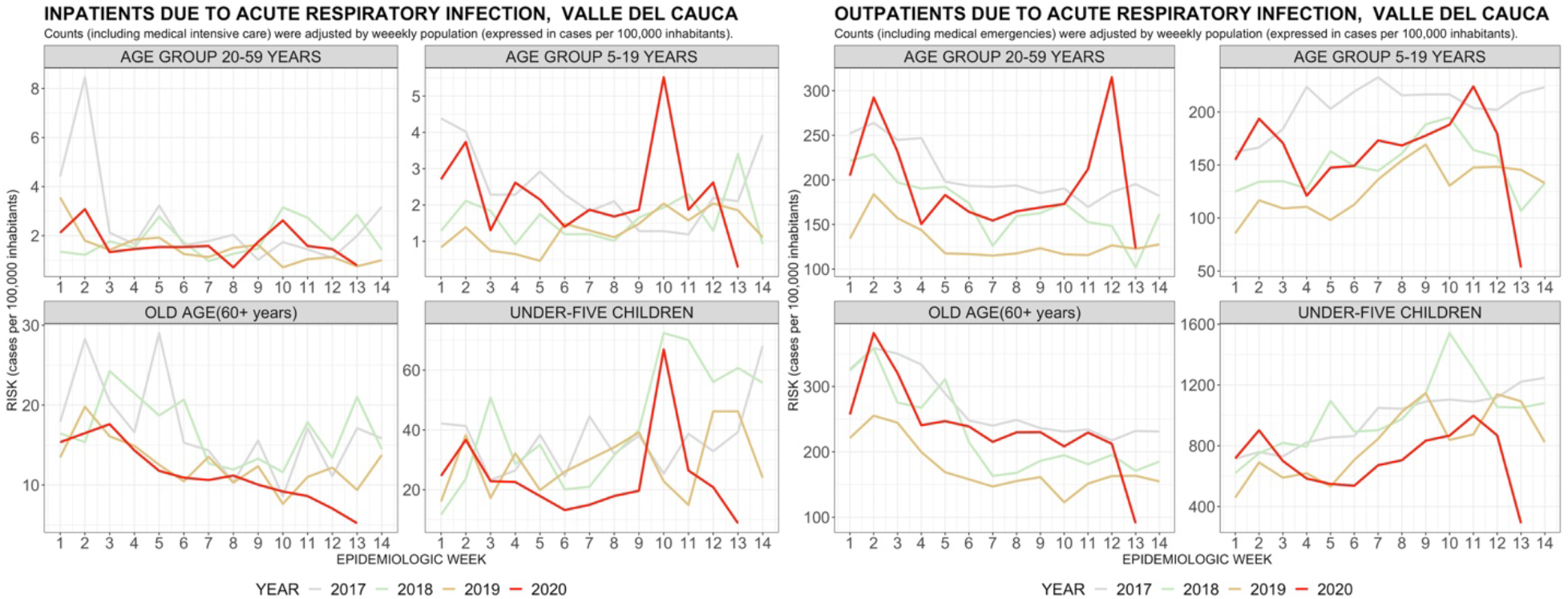
Direct estimation of risk due to Acute Respiratory Infection – ARI, Huila.

**Source:** own calculations.

**Figure 5.**
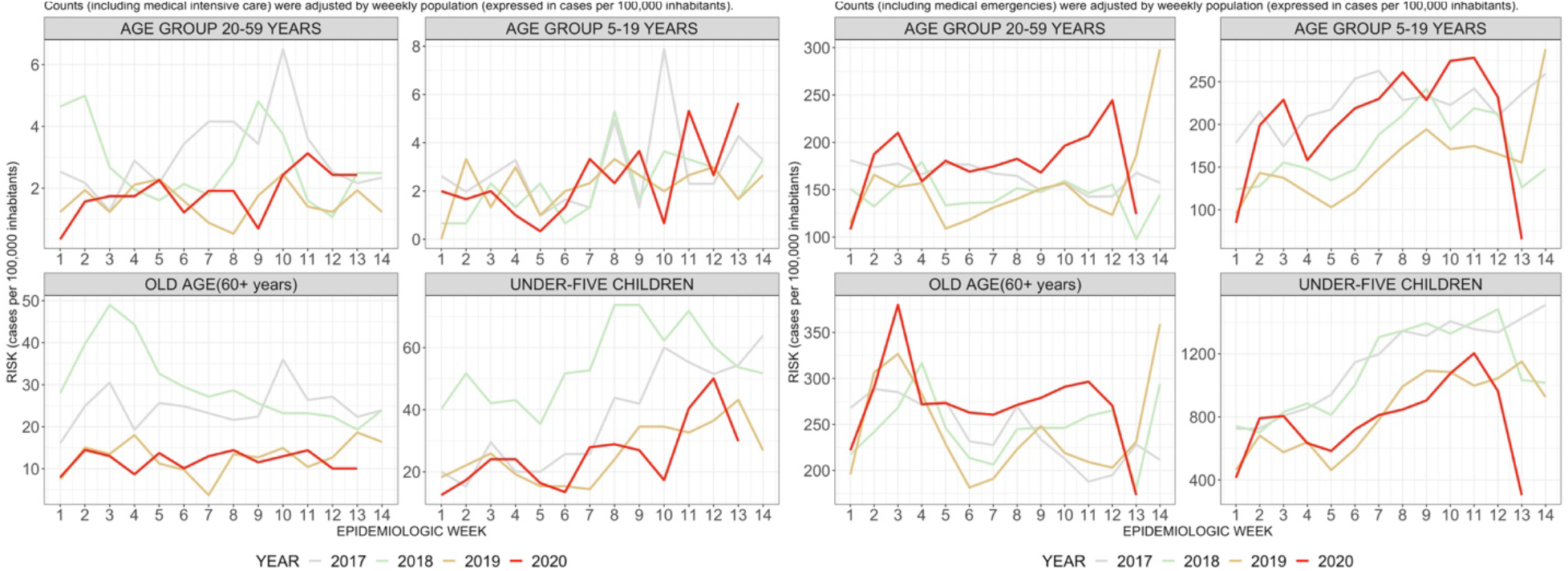
Direct estimation of risk due to Acute Respiratory Infection – ARI, Valle del Cauca.

**Source:** own calculations.

The adjusted Bayesian model presents a good quality of the estimates obtained (see table 2). From the analysis of the symmetric Mean Absolute Percentage Error (sMAPE), it is found that the age groups of higher risk, under 5 years and over 60 years show a percentage deviation of less than 10%, except for the model for Huila of hospital patients that rises to 26%. In general, the models perform well, and the best fit is for inpatient and outpatient cases for children under five which is a risk group of interest.

**Table 2.** Symmetric Mean Absolute Percentage Error (sMAPE) for the quality of outpatient (O) and inpatient (I) model estimates by age group.

|  | Antioquia |  | Bogotá |  | Boyacá |  | Huila |  | Valle |  |
| --- | --- | --- | --- | --- | --- | --- | --- | --- | --- | --- |
| Age group | sMAPE(O)<br>(%) | sMAPE(I)<br>(%) | sMAPE(O)<br>(%) | sMAPE(I)<br>(%) | sMAPE(O)<br>(%) | sMAPE(I)<br>(%) | sMAPE(O)<br>(%) | sMAPE(I)<br>(%) | sMAPE(O)<br>(%) | sMAPE(I)<br>(%) |
| <5 | 4,48 | 8,5 | 3,65 | 4,57 | 4,37 | 8,42 | 6,77 | 15 | 4,45 | 10,6 |
| 5-19 | 6,26 | 13,6 | 7,29 | 7,5 | 6,8 | 14,3 | 8,73 | 28,9 | 6,91 | 14,9 |
| 20-59 | 4,95 | 10 | 5,04 | 6,36 | 5,3 | 11,2 | 8,32 | 25,8 | 4,93 | 12,8 |
| 60+ | 4,89 | 7,63 | 4,58 | 5,46 | 5,26 | 8,51 | 9,72 | 26,3 | 4,67 | 8,07 |

**Source:** own calculations.

The outcome of the Bayesian negative binomial response model in 2020 for the mentioned departments is shown in Figures 6 to 10, in which the 95% credibility interval for the follow-up of these ARI cases that seem striking in the descriptive analysis is presented as red dots in the graphs. In fact, by weeks 11 and 12 in the 5 departments in the 20-59 age group, the observed values of outpatients were outside the range of values expected by the warning at week 10 of the first case of COVID-19 in Colombia, which could have generated a generalized panic that led people to seek medical attention for related symptoms. However, this effect diminished when mobility restrictions were enacted as outpatient cases dropped for all departments in almost all age groups. In fact, by week 13 (lockdown) the number of outpatient cases and hospitalizations of children under five years old for Bogotá and Valle is lower than expected. For Huila, there is an increasing trend of hospitalization cases for the 5-19 year old age group and they are shown as outliers in Figure 9 But in general, there is a drop in outpatient consultations by week 13.

**Figure 6.**
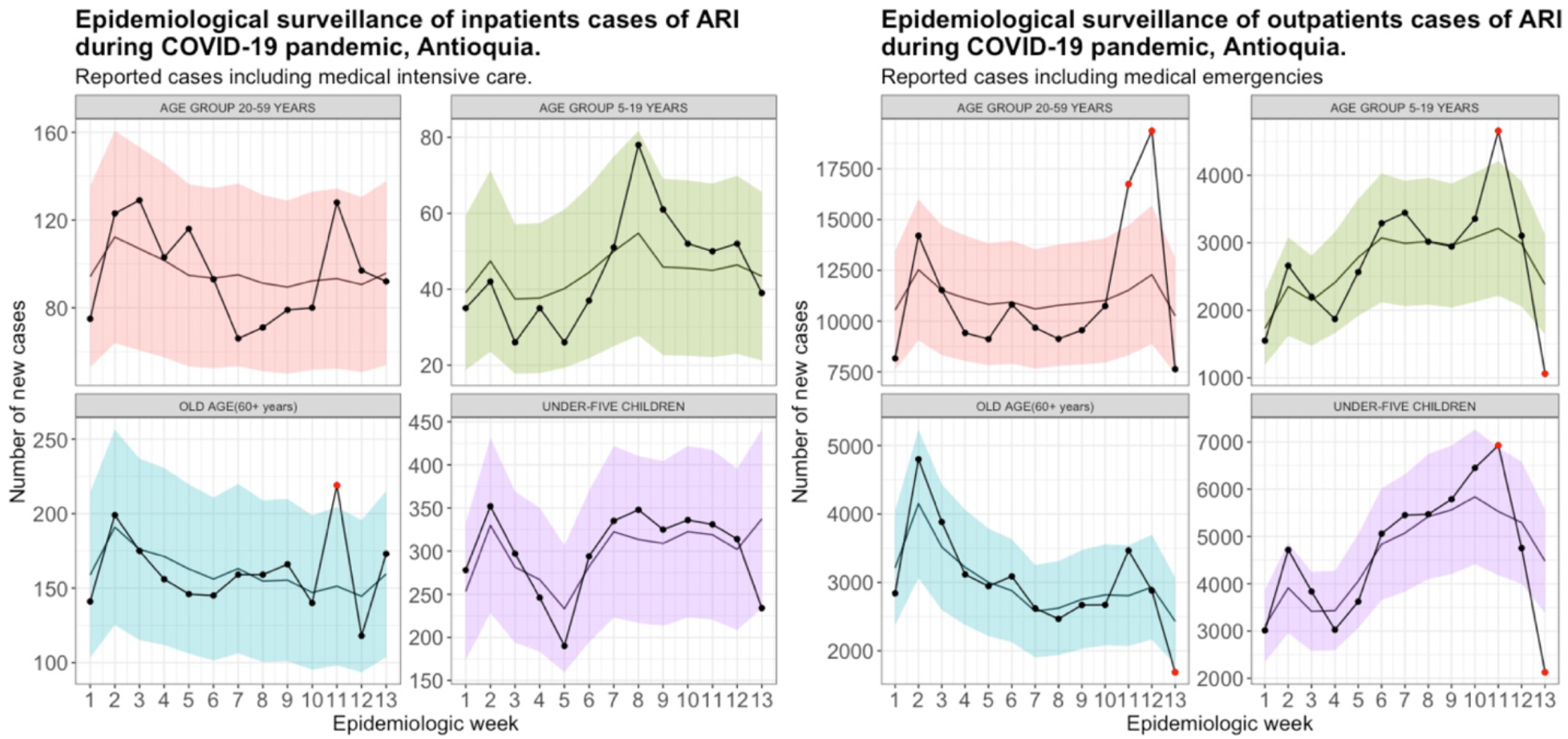
95% credible interval for surveillance of ARI cases during COVID-19 pandemic, Antioquia.

**Source:** own calculations.

**Figure 7.**
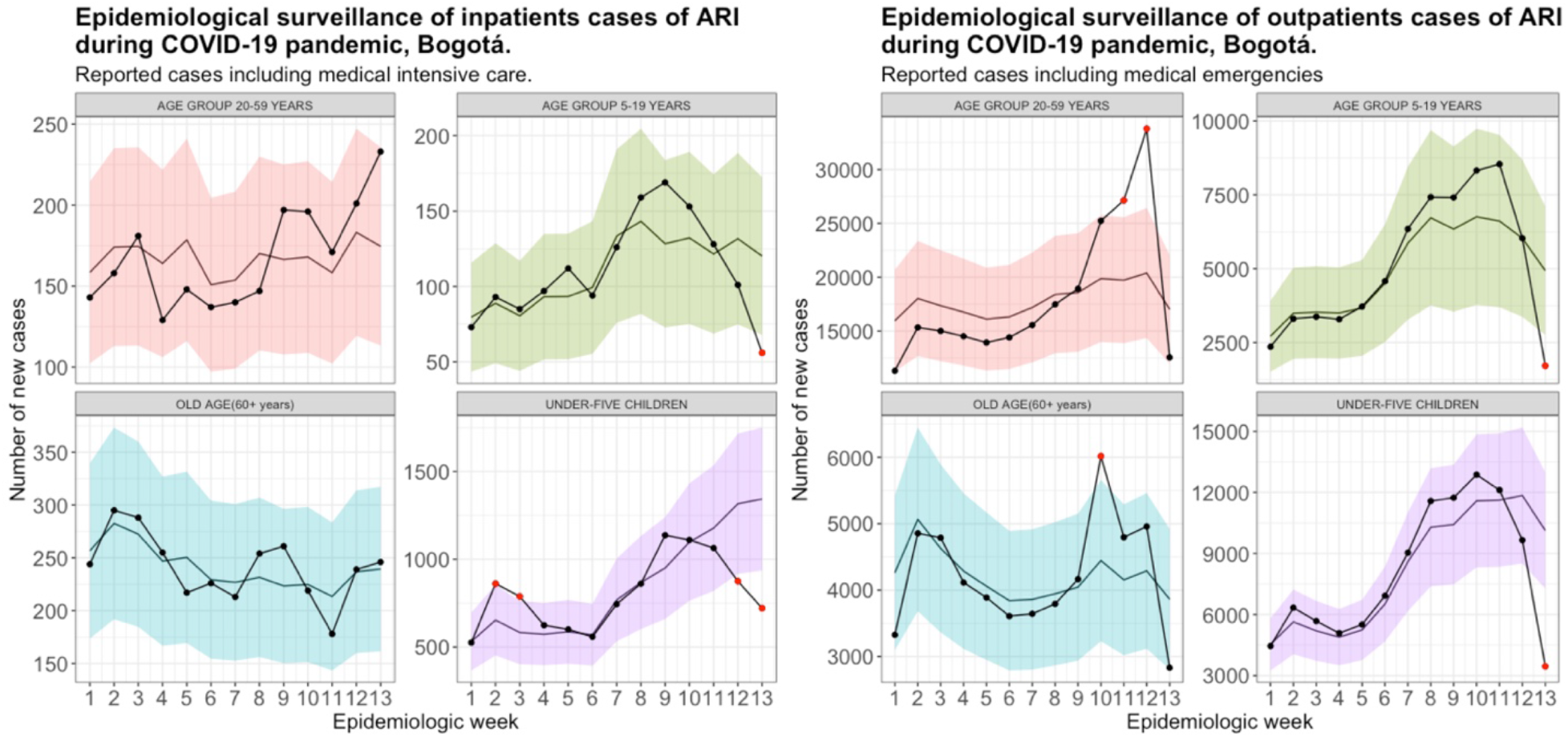
95% credible interval for surveillance of ARI cases during COVID-19 pandemic, Bogotá.

**Source:** own calculations.

**Figure 8.**
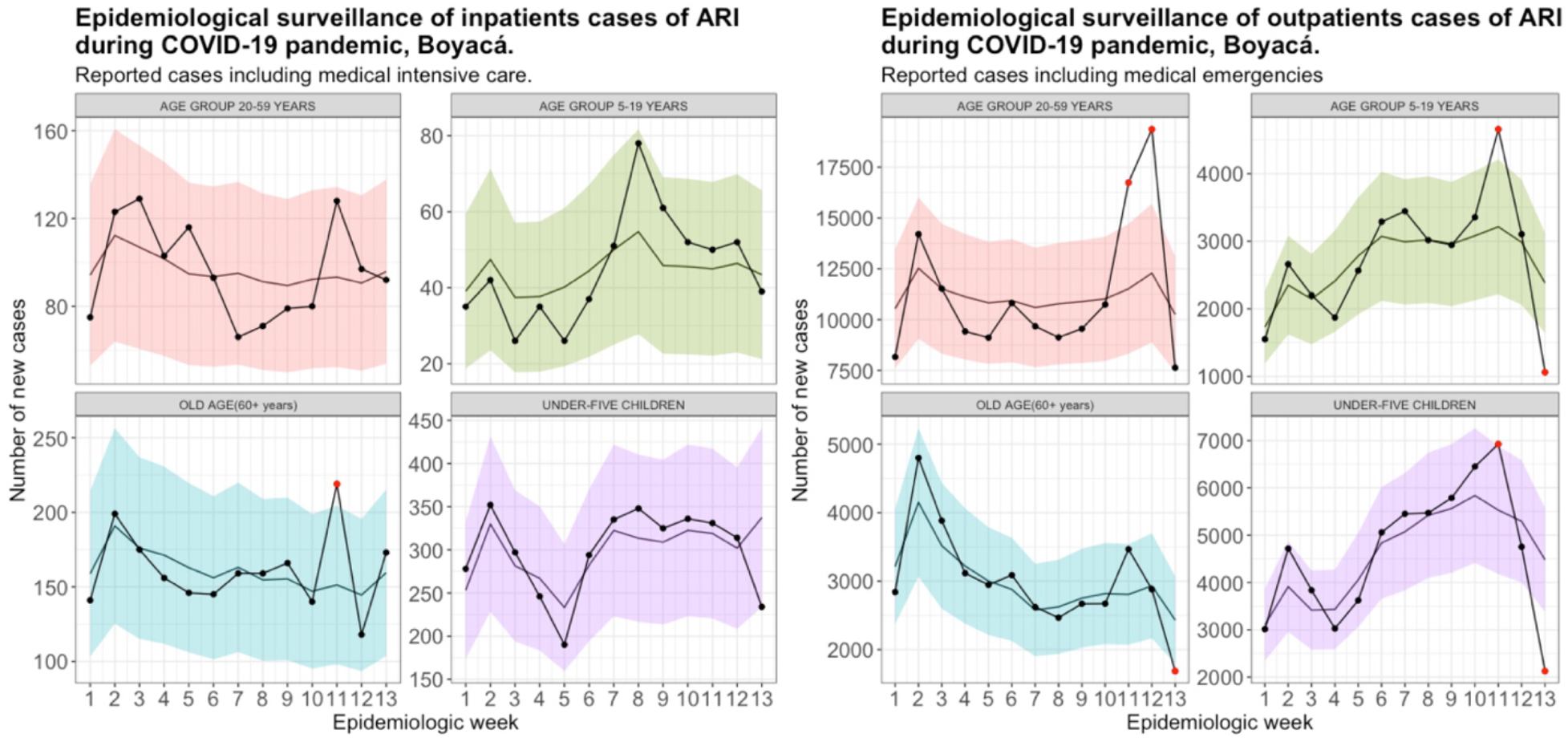
95% credible interval for surveillance of ARI cases during COVID-19 pandemic, Boyacá.

**Source:** own calculations.

**Figure 9.**
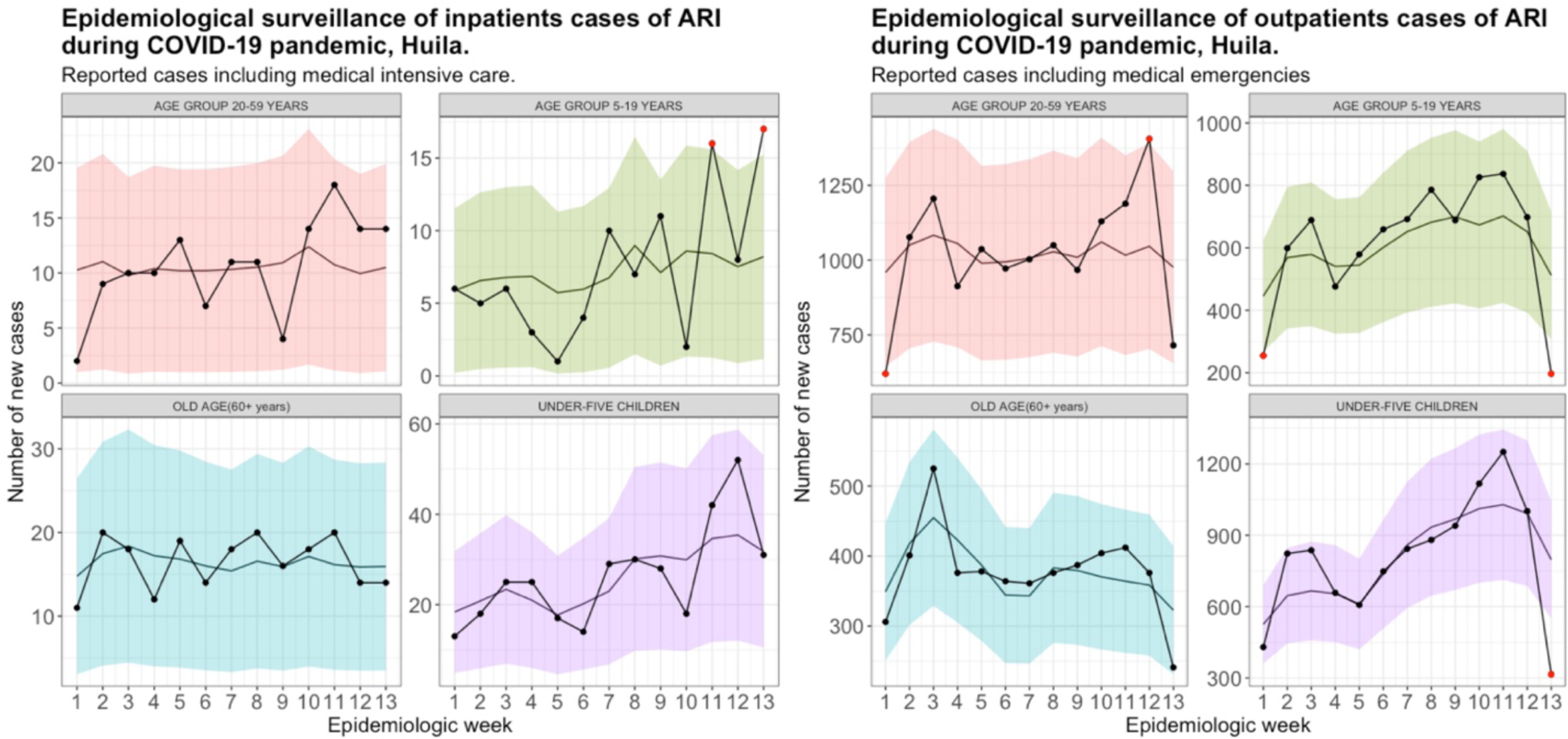
95% credible interval for surveillance of ARI cases during COVID-19 pandemic, Huila.

**Source:** own calculations.

**Figure 10.**
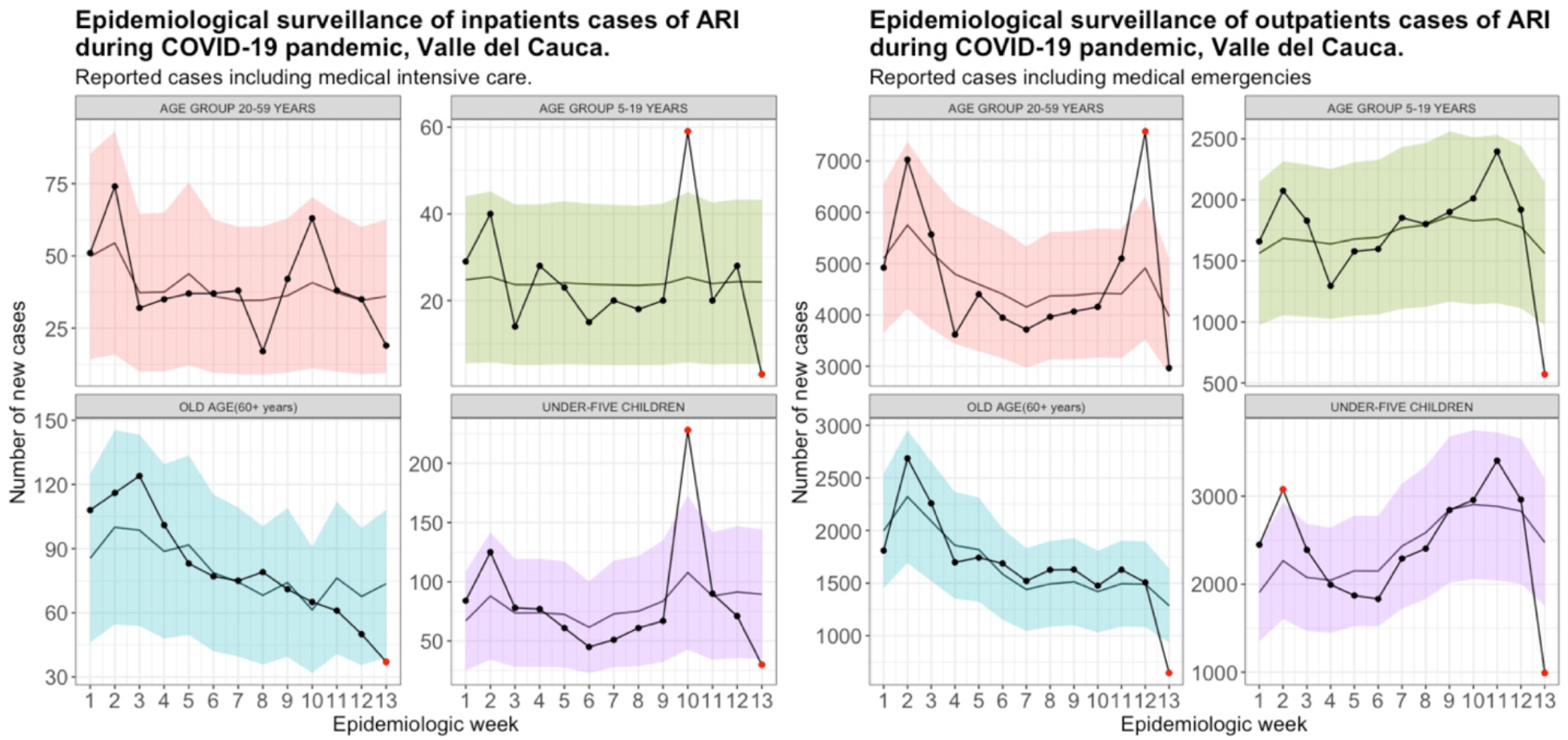
95% credible interval for surveillance of ARI cases during COVID-19 pandemic, Valle del Cauca.

## Conclusions

Although the descriptive analysis is adequate to find atypical patterns in the reported cases of Acute Respiratory Infection in Colombia, the Bayesian model presented allows for the timely detection of those cases that present unexpected behavior since they fall outside the 95% credibility interval estimated from information from epidemiological weeks of previous years, which allows for the evaluation of causes and decision making. Considering the negative binomial distribution that deals with the problem of overdispersion and that is used for discrete variables that represent counts makes the inference reliable and that in places with deficit in the report, it is possible to have a tool to evaluate atypical patterns in the series, without considering the assumption of normal distribution that is used in some surveillance methods and that can lead to biases. As a method of surveillance during the pandemic, the result presents sufficient elements for the evaluation of those rare cases that require assessment and allow intervention and control in health systems.

## Data Availability

The data are not available because they are sensitive health data, and are protected by
Colombia's habeas data law (Ley 1581 de 2012).

## Footnotes

1 Makridakis (6) shows the benefits of this index, which is one of the most used in the international literature on forecast calculations.

2 When descriptively analyzing the risk of the 33 departments of Colombia, these 5 were the ones that exposed the temporal trends of greatest alarm (abrupt drop in health care).

